# Exploring the Accuracy of Differentiation-Based Regressive Models in Disease Forecasting

**DOI:** 10.1101/2023.10.26.23297654

**Authors:** Rojina Karimirad

## Abstract

Predictive models have been able to foresee outbreaks of mosquito-borne diseases such as malaria and map Ebola outbreaks^1^. This has allowed health organizations to plan the amount of resources and the number of healthcare workers needed more effectively, on top of finding out other useful data such as the locations most vulnerable to the disease and the demographics most affected. It can therefore be assumed that predictive analytics can reduce the amount of economic and non-economic burden caused by other epidemics as well, with COVID-19 being an obvious example.

---

To explore the use of predictive analytics in disease forecasting and in COVID-19 specifically, I decided to test the accuracy of a differentiation-based regression model on data provided by the Ontario Data Catalogue^2^ and then compare its performance to other methods of calculating regression. To make the prediction more personal, I decided to use data pertaining to the closest Ontario Health region to me, which is Central Ontario. The original set of data provided the daily number of hospitalizations since the beginning of the virus outbreak, however the data belonging to the year 2020 was discarded due to the assumption that the overwhelming surge to hospitals at the beginning of the pandemic would skew the data and hence the regression model. The reduced raw set of data covers COVID-19 cases in the hospital from January 1, 2021 to December 31, 2022, where the date is the independent variable, and the number of hospitalizations is the dependent variable. It can be found in *Appendix A*. To clearly display the data spanning two years on a single table, the number of hospitalizations for each ten days in the data were put into one group, and to process the data, a numerical value was assigned to each ten-day group, so January 1, 2021 to January 10, 2021 was assigned 1, January 11 to January 20 was assigned 2, etc. Since there are 730 days in two years, there ended up being 73 groups of 10 days in total. The new data table can be seen in *Appendix B*.

The scatterplot showing the number of hospitalizations due to COVID-19 in the years 2021 and 2022 and their corresponding ten-day groups is shown in *Graph 1*.

**Graph 1:**
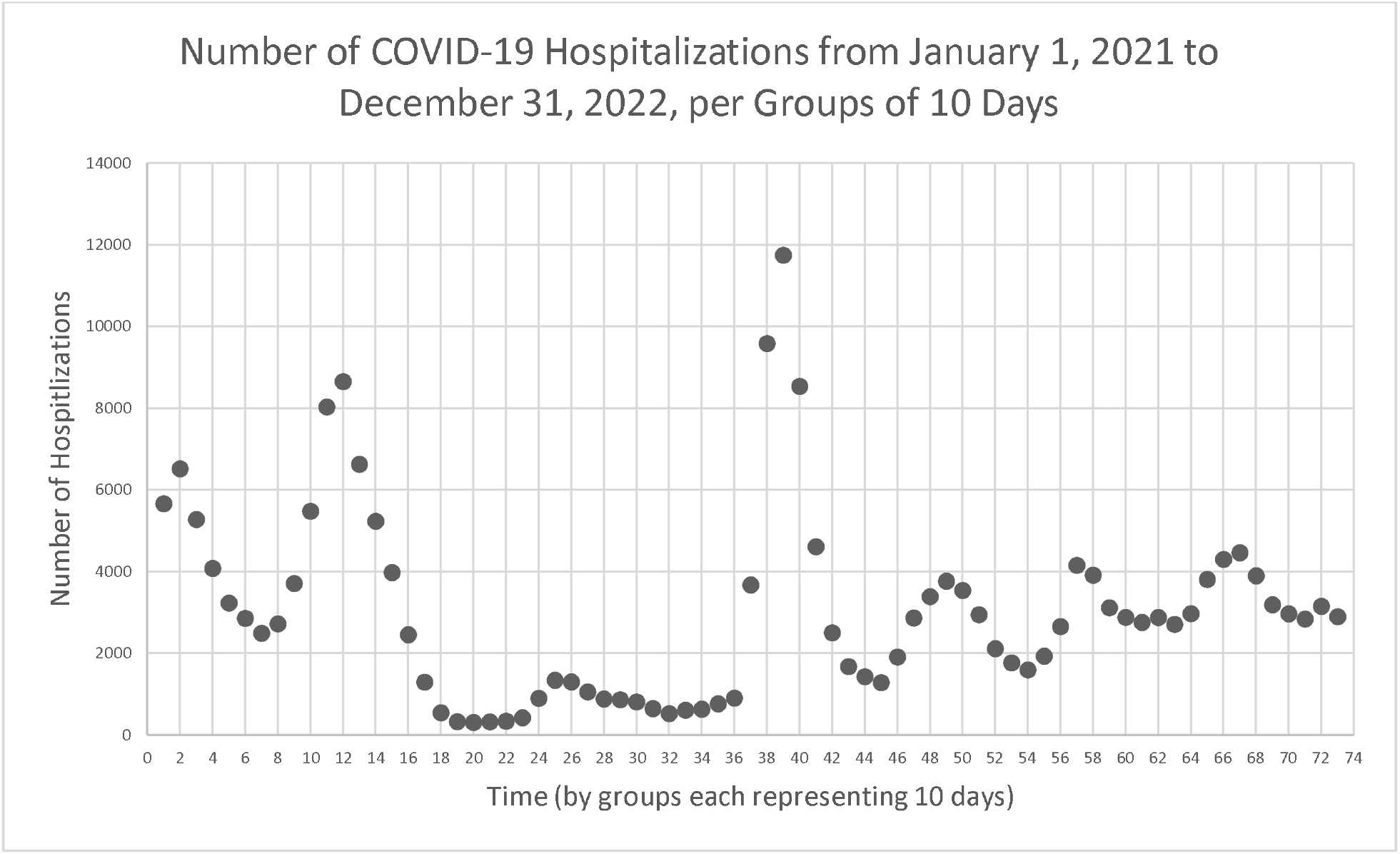
Time series plot showing the number of COVID-19 hospitalizations in 2021 and 2022 in groups of 10 days

By only looking at the scatterplot, we notice certain outliers within the data. Outliers can negatively impact the accuracy of a regression model, so their elimination would be beneficial. Since the independent variable of the data is groups of ten days, a unit of time, the data can be categorized as a time series and the above scatterplot can be considered a time series plot. This makes us able to use methods typically utilized for single-variable data, such as the interquartile range, quartile values, and the lower and upper inner fences of the number of hospitalizations to calculate the outliers, because it is impossible for the *x* or time values that are consistently increasing by 1 group or 10 days to produce outliers on their own^3^.

The lower and upper inner fences of the dataset can be used to find the set’s outliers, with any value that lies beyond these two points being an outlier. Since the formulae for the lower and upper fences are, respectively:

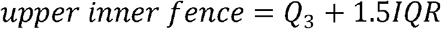

Where *Q*_*3*_ is Quartile 3 and *IQR* is the interquartile range.

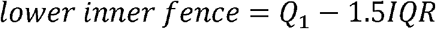

Where *Q*_*1*_ is Quartile 1 and *IQR* is the interquartile range.

And the formula for interquartile range or *IQR* is:

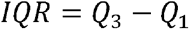

Where *IQR* is the interquartile range, *Q*_*1*_ is Quartile 1 and *Q*_*3*_ is Quartile 3.

The values for Quartiles 1 and 3 need to be calculated. For the quartile values to be determined, the number of hospitalizations for each group of 10 days were placed in an increasing order and assigned term numbers based on their place in the newly ordered list, as shown in *Appendix C*. The formula for calculating the term number for the value of Quartile 1 is,

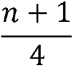

Where *n* is the number of terms, which in this case is 73.

Substituting *n* = 73 into this formula, we get:

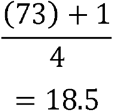

Since there is no 18.5^th^ term, the mean of the values of the 18^th^ and 19^th^ terms, which are obtained from *Appendix C*, is used to determine *Q*_*1*_.

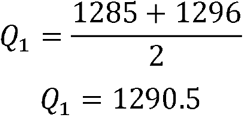

The time values of Quartile 3 can be calculated using a similar formula,

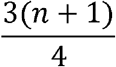

Where *n* is the number of terms.

Substituting *n* = 73 once again,

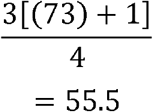

Again, since there is no 55.5^th^ term, the mean of the 55^th^ and 56^th^ terms’ values from *Appendix C* is used to calculate *Q*_*3*_.

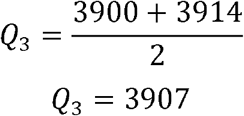

Having calculated Quartiles 1 and 3, the values can be substituted into the previously stated formula for interquartile range,

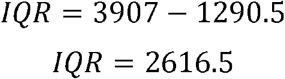

The interquartile range, along with *Q*_*1*_ and *Q*_*3*_, can then be used to find the upper inner fence, as shown below,

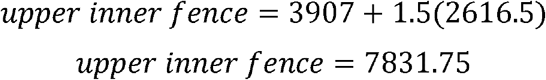

And similarly, the lower inner fence,

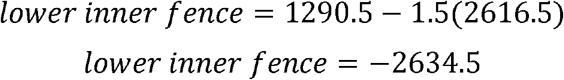

Since the lower inner fence was calculated to be a negative number, and the number of hospitalizations cannot be negative, it can be concluded that there are no *y* values in the time series that are outliers due to being too small.

The upper inner fence, however, provides a limit for how large the values for the number of hospitalizations can be without skewing the data and therefore the regression model that will be produced. The following groups and their corresponding values for number of hospitalizations were taken from *Appendix B* and noted as outliers on the basis of being larger than the upper inner fence, 7831.75:

The outliers were removed from the dataset and replaced with the means of the two values before and after each of them in *Appendix B*, to stop them from impacting the accuracy of the regression, as shown in the following table:

The data excluding the outliers and instead including their newly assigned values, which will be used for the regression model, is shown in *Appendix D*. The graph visualizing the new data on the same scale can be seen below.

After having removed the outliers from the data, we have a dataset that would produce a more reliable regression model. There however needs to be a method of checking the accuracy of the regression model, which is where Test Train Split will be used. Test Train Split is a model validation procedure that checks the accuracy of a regression model’s performance on new data through interpolation and the data already available^4^. The Split refers to the split of the data into Train, which is 80% of the total data and will be used to calculate the regression equation, and Test, which is the remaining 20% and will be used to test the accuracy of the regression model. Since 80% of 73, the total number of data points, is 58.4 and not a whole number, it is rounded to 58. Similarly, 20% of 73, 14.6, is rounded to 15. The fifteen numbers that will only be used to test for the accuracy of and not to come up with the regression equation were randomized using a Java program I coded myself, linked in *Appendix E*. The program randomly printed the *x* values that can be seen in *Table 3* with their corresponding *y* values:

**Table 1:** Outliers determined based on the upper inner fence value of 7831.75.

| **Group** | 11 | 12 | 38 | 39 | 40 |
| --- | --- | --- | --- | --- | --- |
| **Number of Hospitalizations** | 8033 | 8654 | 9586 | 11749 | 8538 |

**Table 2:** Outliers replaced by the mean of the inlier values nearest to them.

| **Group** | 11 | 12 | 38 | 39 | 40 |
| --- | --- | --- | --- | --- | --- |
| **New Number of Hospitalizations** | 6055.5 | 6055.5 | 4143 | 4143 | 4143 |

**Table 3:** Fifteen randomly generated values making up the Test split, in increasing order of time.

| Time ($x$) | Number of Hospitalizations ($y$) | Time ($x$) | Number of Hospitalizations ($y$) |
| --- | --- | --- | --- |
| 5 | 3232 | 46 | 1912 |
| 8 | 2723 | 51 | 2947 |
| 17 | 1296 | 56 | 2656 |

<sup>4</sup> Michael Galarnyk, "Understanding Train Test Split," Built In, July 28, 2022, https://builtin.com/data-science/train-test-split.

|  |  |  |  |
| --- | --- | --- | --- |
| 26 | 1302 | 64 | 2971 |
| 30 | 812 | 69 | 3190 |
| 35 | 766 | 71 | 2841 |
| 37 | 3675 | 72 | 3153 |
| 43 | 1677 |  |  |

The final version of the processed data, excluding the outliers and only containing the Train split, can be seen in *Appendix F*.

To come up with the most accurate regression equation for this data, we can use the concept of the loss or cost function. The loss function is a measure of how badly a regression model can estimate the relationship between *x* and *y*, and it can be written using sigma notation, signifying summation^5^. The way the loss function measures the performance of the model is by calculating the distance between the expected versus real value of *y* at *x*, with *x* and *y* being the group and number of hospitalization values recorded in *Appendix F*. The loss function for linear regression is written as:

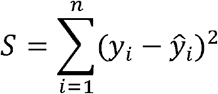

Where *S* is the loss function, *ŷ*_*i*_ is the expected *i*th value of *y*, and *y*_*i*_ is the actual *i*th value of *y*.

The difference between the expected and actual value of *y* is squared to avoid negative error values. This issue could also be avoided via finding the absolute value of the difference, however that would make the function indifferentiable at some points, which would make us unable to minimize the error using derivatives. Squaring the error also further penalizes the regression model for making errors, as it would make a small error, like one by 20 units, appear as 400 instead.

Now, the goal is to find the regression model that achieves the lowest possible amount of loss. To do this, we need to identify the unknown coefficients and constants in the equation of a linear regression model, which is:

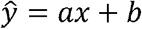

Where *a* is the slope of the regression model, or the coefficient of *x*, and *b* is the y-intercept, or the constant.

Substituting the equation of the regression model into the loss function, we get:

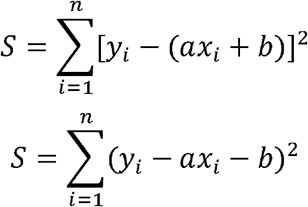

To find the values of *a* and *b* that would minimize the amount of loss, we need to partially differentiate the loss function with respect to the two unknowns. We can start with *b*, using the chain rule,

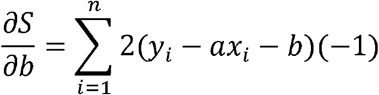

To minimize the value of *b* and to find the “critical numbers” of the loss function with respect to *b*, we set the partial derivative to 0 and isolate for *b*:

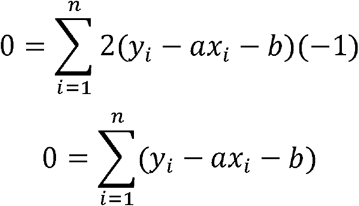

Breaking the summation up and factoring out *a*,

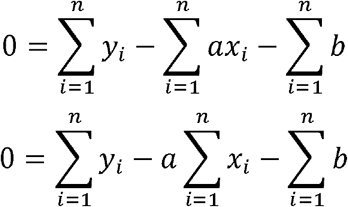

Solving for 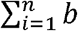 with respect to *n*,

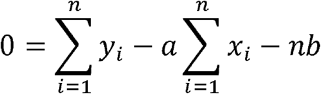

Adding both sides by *nb*,

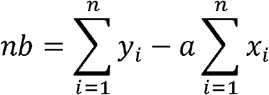

Dividing both sides by *n*,

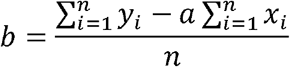

Looking at the resulting equation closely, we notice that the summation of *y* values divided by *n*, which is the number of terms, is equal to the mean of *y* values, or 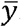. The same can be said for the summation of *x* values divided by *n*, which is equal to 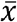.

Substituting in 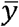 and 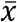, we get,

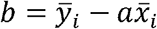

We will leave *b* for now, and partially differentiate the loss function with respect to *a* this time:

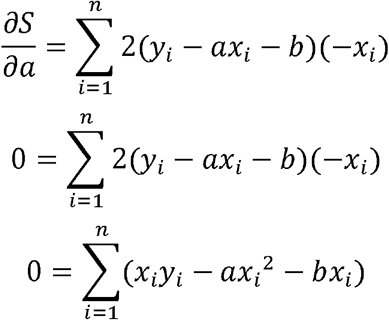

Substituting in 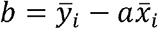,

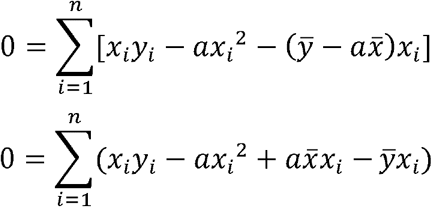

Isolating *a*,

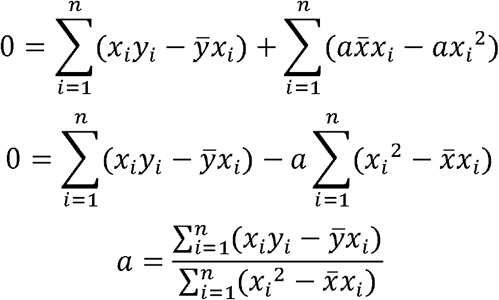

Having found the equations for both *a* and *b*, the means of the *x* and *y* values from *Appendix F* were found to solve for *a* and *b*, using the following formula,

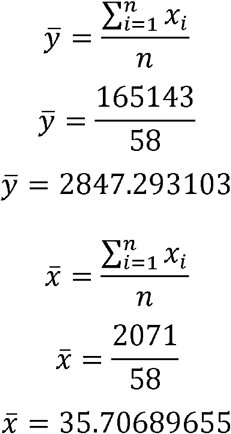

The mean values and the values for *x* and *y* obtained from *Appendix F* were substituted into *a*.

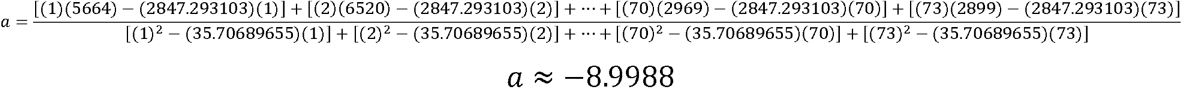

*a* was rounded to five significant digits. Substituting *a* into the equation of *b*,

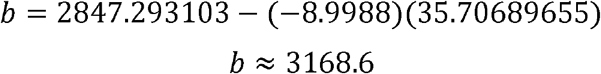

*b* was also rounded to five significant digits. Substituting the values of *a* and *b* into the equation for line of best fit,

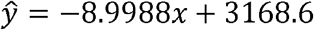

Having found the equation of the linear regression model, we can graph the time series plot representing the data along with the regression. The fifteen Test values are also on the graph, represented by a different shade of grey to signify that they did not influence the regression line.

Visually, the regression seems to pass through some of the Test points and be far away from others. The difference between the actual Test points versus the ones predicted by the regression can be found by subtracting the number of hospitalizations of each Test point by the value obtained when substituting their time values into the regression equation and taking the absolute value of the difference. A sample calculation of this is shown below for the first Test point at *x* = 5,

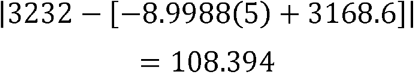

The sum of the differences can then be divided by 15, the total number of Test points, to find the Mean Absolute Error of the regression model^6^. This process is shown in *Table 4*.

**Table 4:** Test Train Split calculations to determine the Mean Absolute Error and evaluate the accuracy of the regression model.

| Time (x) | Actual Number of Hospitalizations (y) | Expected Number of Hospitalizations ($\hat{y}$) | Absolute Error (Difference between Actual and Expected Test Values) | Mean Absolute Error |
| --- | --- | --- | --- | --- |
| 5 | 3232 | 3123.606 | 108.394 | 867.0314 |
| 8 | 2723 | 3096.6096 | 373.6096 |  |
| 17 | 1296 | 3015.6204 | 1719.6204 |  |
| 26 | 1302 | 2934.6312 | 1632.6312 |  |
| 30 | 812 | 2898.636 | 2086.636 |  |
| 35 | 766 | 2853.642 | 2087.642 |  |
| 37 | 3675 | 2835.6444 | 839.3556 |  |
| 43 | 1677 | 2781.6516 | 1104.6516 |  |
| 46 | 1912 | 2754.6552 | 842.6552 |  |
| 51 | 2947 | 2709.6612 | 237.3388 |  |
| 56 | 2656 | 2664.6672 | 8.6672 |  |
| 64 | 2971 | 2592.6768 | 378.3232 |  |
| 69 | 3190 | 2547.6828 | 642.3172 |  |
| 71 | 2841 | 2529.6852 | 311.3148 |  |
| 72 | 3153 | 2520.6864 | 632.3136 |  |

A Mean Absolute Error of 867.0314 is high for a dataset with numbers that range from 309 to 6630, hinting at the regression model not being a good fit for the data. This result led to me looking back at my process and attempting to identify limitations that caused the calculated regression model to not be well representative of the data.

The main limitation I found was the shape of the regression model. The loss function I optimized minimized the inaccuracy of a linear regression model, but data pertaining to a pandemic may not have a linear trend as the rate of the drop in the number of hospitalizations decreases overtime as the total number of cases decreases. Data of such nature can be represented by a logarithmic or polynomial regression model. As an extension, the accuracy of the calculated linear regression model versus logarithmic and polynomial regression models can be compared via the *R*^*2*^ value or the coefficient of determination. The *R*^*2*^ value is a value from 0 to 1, with 0 being the least accurate and 1 being the most accurate, that is calculated based on the ratio of the residual sum of squares, which measures the deviation between the actual data and the data predicted by the regression model, to the total sum of squares, which is the deviation between the actual data and the mean^7^. The formula for the *R*^*2*^ value, therefore, is:

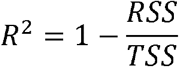

Where *RSS* is the residual sum of squares and *TSS* is the total sum of squares.

It is important to note that the accuracy of linear regression is typically not measured using the *R*^*2*^ value and is instead determined based on the *r* value, or the correlation coefficient. However, for the sake of comparing a linear regression model with non-linear models, the *R*^*2*^ value will be used.

Below is a graph containing the same data as *Graph 3*, but instead with a logarithmic regression curve and its *R*^*2*^ value generated via Excel.

**Graph 2:**
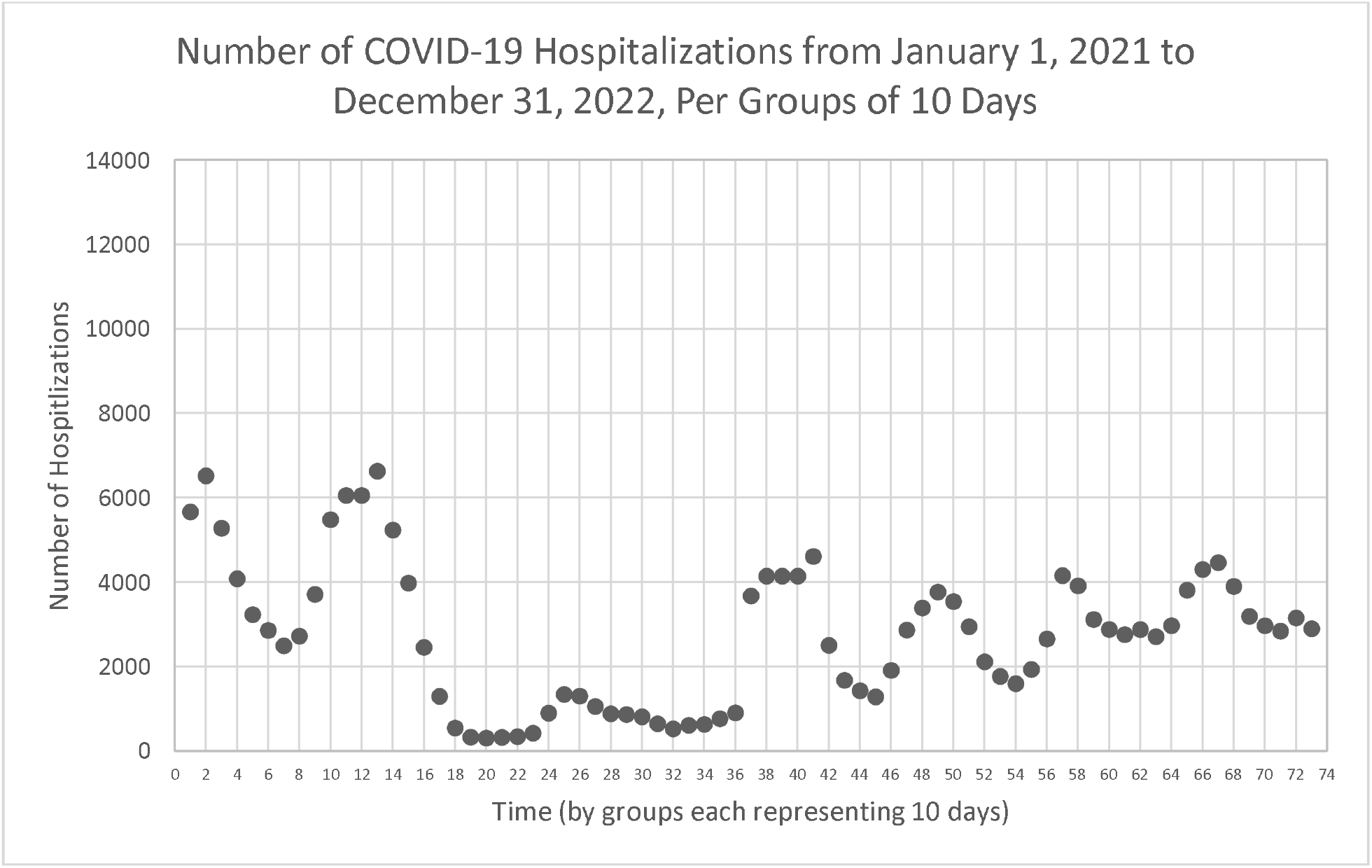
Time series plot showing the number of COVID-19 hospitalizations in 2021-2022 in groups of 10 days, without outliers

**Graph 3:**
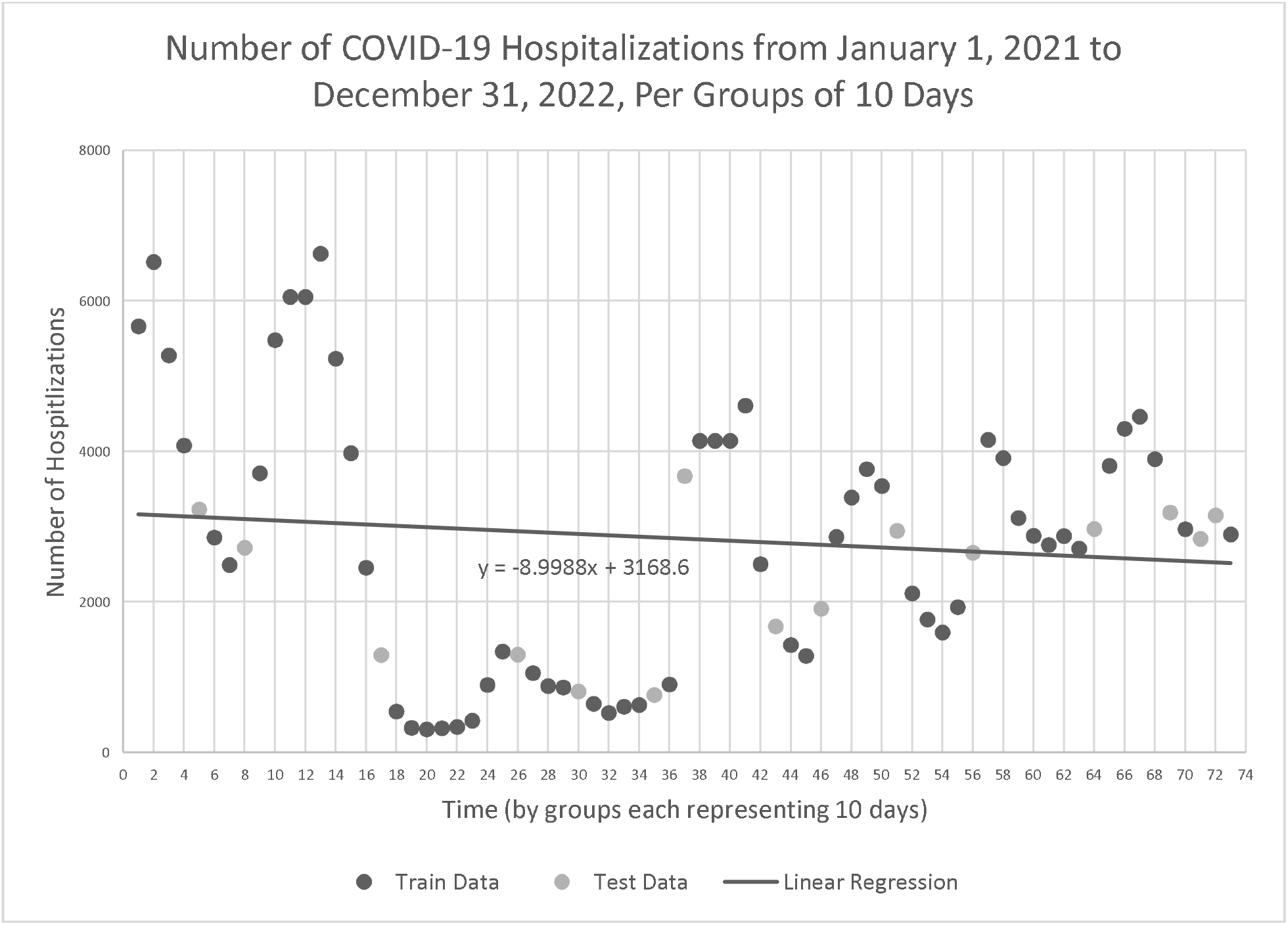
Time series plot showing the number of COVID-19 hospitalizations in 2021-2022 in groups of 10 days, including the linear regression model, separated into the Test and Train splits

The *R*^*2*^ value of the linear regression drawn in *Graph 3* was calculated to be 0.011, again via Excel. The *R*^*2*^ value of the logarithmic regression model seen on *Graph 4*, 0.1115, is around ten times greater than 0.011, confirming that my identification of the biggest limitation being the shape was correct and showing that a logarithmic regression would fit the data better.

**Graph 4:**
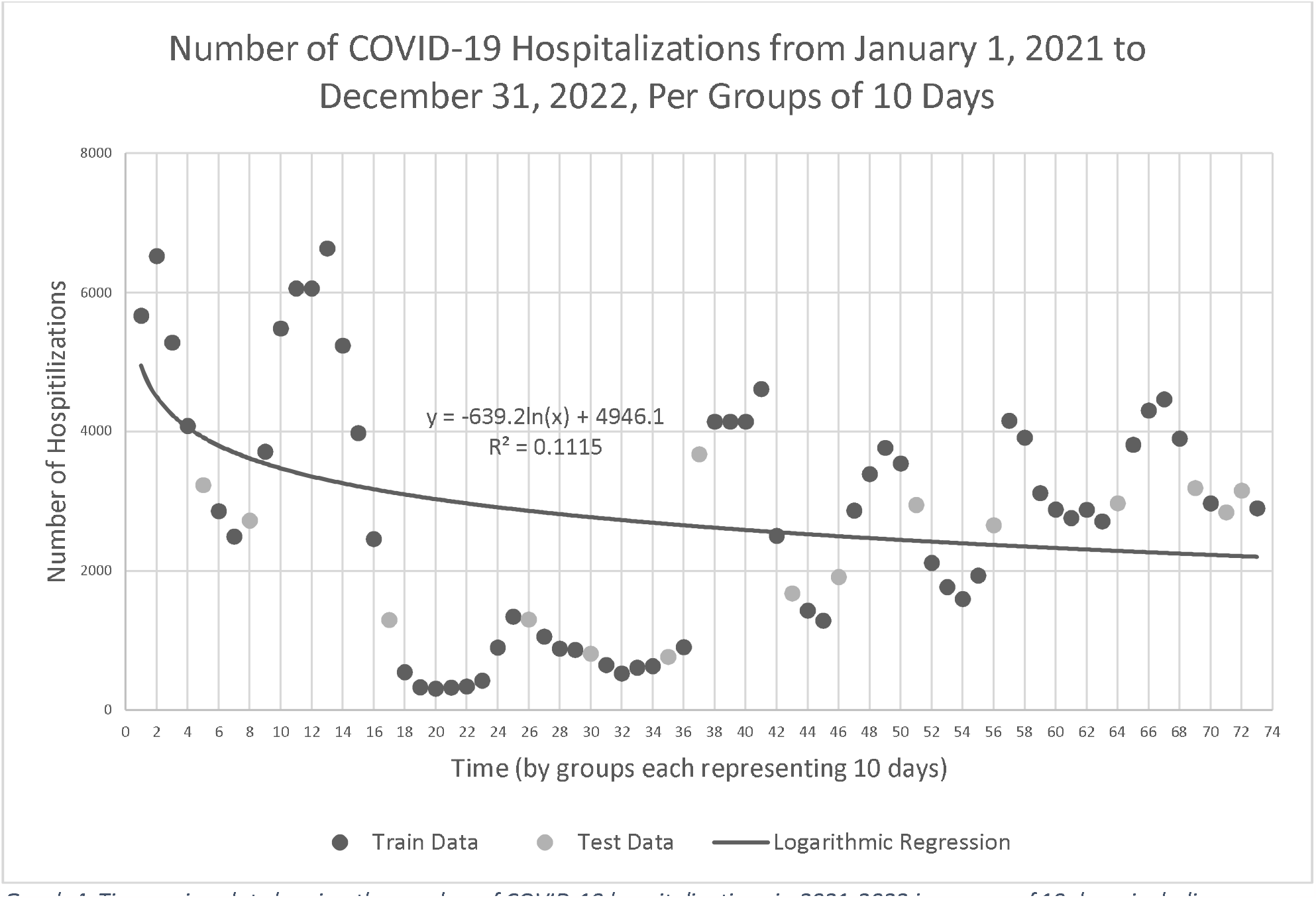
Time series plot showing the number of COVID-19 hospitalizations in 2021-2022 in groups of 10 days, including a logarithmic regression model, separated into the Test and Train splits

**Graph 5:**
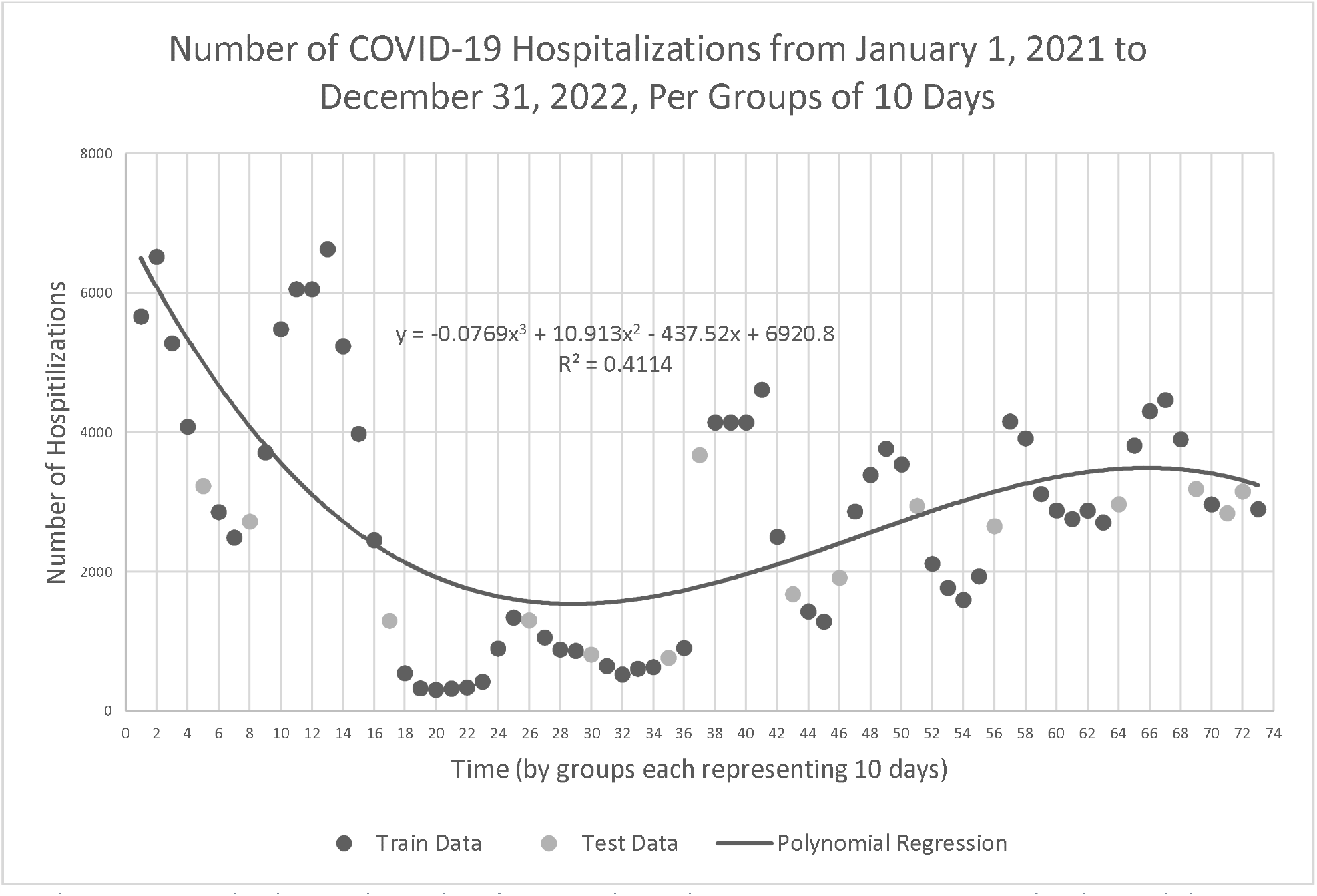
Time series plot showing the number of COVID-19 hospitalizations in 2021-2022 in groups of 10 days, including a polynomial regression model, separated into the Test and Train splits

The polynomial regression model and its *R*^*2*^ value were also found using Excel and can be seen in the graph below.

The *R*^*2*^ value of the polynomial regression model, 0.4114, is even higher, being around four times greater than that of the logarithmic regression model and around forty times greater than that of the linear model. This proves that the shape of the model was in fact the issue with the lack of inaccuracy shown by Test Train Split in *Table 4*.

It is important to deduce the reason for the linear regression model’s inaccuracy to answer my original research question: can differentiation-based regressive models provide accurate disease forecasting? The answer is not no, because the limitation was confirmed to be the linear model’s shape and not the method by which its equation was found. Since differentiation was used to minimize the error calculated by the loss function, we can be certain that the derived linear equation was the best possible linear model for the data. So, as an even further extension, if differentiation-based regression was applied to non-linear regression, it could absolutely be used to forecast the progression of diseases such as COVID-19.

## Data Availability

All data produced are available online at

https://data.ontario.ca/dataset/covid-19-cases-in-hospital-and-icu-by-ontario-health-region

---

**Appendix A:** Raw data from January 1, 2021 to December 31, 2022, obtained from the Ontario Data Catalogue

| Date | Number of Hospitalizations | Date | Number of Hospitalizations | Date | Number of Hospitalizations | Date | Number of Hospitalizations | Date | Number of Hospitalizations |
| --- | --- | --- | --- | --- | --- | --- | --- | --- | --- |
| 2021-01-01 | 517 | 2021-03-04 | 250 | 2021-05-05 | 705 | 2021-07-06 | 30 | 2021-09-06 | 134 |
| 2021-01-02 | 503 | 2021-03-05 | 249 | 2021-05-06 | 661 | 2021-07-07 | 30 | 2021-09-07 | 134 |
| 2021-01-03 | 534 | 2021-03-06 | 233 | 2021-05-07 | 632 | 2021-07-08 | 32 | 2021-09-08 | 137 |
| 2021-01-04 | 542 | 2021-03-07 | 237 | 2021-05-08 | 592 | 2021-07-09 | 36 | 2021-09-09 | 132 |
| 2021-01-05 | 562 | 2021-03-08 | 245 | 2021-05-09 | 566 | 2021-07-10 | 33 | 2021-09-10 | 141 |
| 2021-01-06 | 598 | 2021-03-09 | 257 | 2021-05-10 | 552 | 2021-07-11 | 31 | 2021-09-11 | 129 |
| 2021-01-07 | 595 | 2021-03-10 | 256 | 2021-05-11 | 586 | 2021-07-12 | 34 | 2021-09-12 | 126 |
| 2021-01-08 | 592 | 2021-03-11 | 255 | 2021-05-12 | 524 | 2021-07-13 | 37 | 2021-09-13 | 119 |
| 2021-01-09 | 603 | 2021-03-12 | 255 | 2021-05-13 | 530 | 2021-07-14 | 33 | 2021-09-14 | 128 |
| 2021-01-10 | 618 | 2021-03-13 | 264 | 2021-05-14 | 528 | 2021-07-15 | 33 | 2021-09-15 | 124 |
| 2021-01-11 | 636 | 2021-03-14 | 261 | 2021-05-15 | 512 | 2021-07-16 | 33 | 2021-09-16 | 136 |
| 2021-01-12 | 700 | 2021-03-15 | 273 | 2021-05-16 | 518 | 2021-07-17 | 26 | 2021-09-17 | 130 |
| 2021-01-13 | 671 | 2021-03-16 | 271 | 2021-05-17 | 530 | 2021-07-18 | 22 | 2021-09-18 | 116 |
| 2021-01-14 | 664 | 2021-03-17 | 272 | 2021-05-18 | 524 | 2021-07-19 | 27 | 2021-09-19 | 105 |
| 2021-01-15 | 665 | 2021-03-18 | 270 | 2021-05-19 | 506 | 2021-07-20 | 29 | 2021-09-20 | 101 |
| 2021-01-16 | 647 | 2021-03-19 | 278 | 2021-05-20 | 477 | 2021-07-21 | 30 | 2021-09-21 | 107 |
| 2021-01-17 | 624 | 2021-03-20 | 285 | 2021-05-21 | 469 | 2021-07-22 | 25 | 2021-09-22 | 101 |
| 2021-01-18 | 633 | 2021-03-21 | 294 | 2021-05-22 | 442 | 2021-07-23 | 37 | 2021-09-23 | 104 |
| 2021-01-19 | 641 | 2021-03-22 | 317 | 2021-05-23 | 411 | 2021-07-24 | 35 | 2021-09-24 | 105 |
| 2021-01-20 | 639 | 2021-03-23 | 334 | 2021-05-24 | 410 | 2021-07-25 | 33 | 2021-09-25 | 112 |
| 2021-01-21 | 573 | 2021-03-24 | 343 | 2021-05-25 | 423 | 2021-07-26 | 35 | 2021-09-26 | 103 |
| 2021-01-22 | 564 | 2021-03-25 | 334 | 2021-05-26 | 406 | 2021-07-27 | 37 | 2021-09-27 | 103 |
| 2021-01-23 | 566 | 2021-03-26 | 345 | 2021-05-27 | 397 | 2021-07-28 | 33 | 2021-09-28 | 102 |
| 2021-01-24 | 548 | 2021-03-27 | 374 | 2021-05-28 | 379 | 2021-07-29 | 30 | 2021-09-29 | 92 |
| 2021-01-25 | 537 | 2021-03-28 | 375 | 2021-05-29 | 327 | 2021-07-30 | 31 | 2021-09-30 | 89 |
| 2021-01-26 | 550 | 2021-03-29 | 392 | 2021-05-30 | 315 | 2021-07-31 | 28 | 2021-10-01 | 82 |
| 2021-01-27 | 512 | 2021-03-30 | 431 | 2021-05-31 | 304 | 2021-08-01 | 30 | 2021-10-02 | 79 |
| 2021-01-28 | 493 | 2021-03-31 | 467 | 2021-06-01 | 292 | 2021-08-02 | 27 | 2021-10-03 | 79 |
| 2021-01-29 | 469 | 2021-04-01 | 469 | 2021-06-02 | 278 | 2021-08-03 | 30 | 2021-10-04 | 80 |
| 2021-01-30 | 466 | 2021-04-02 | 467 | 2021-06-03 | 270 | 2021-08-04 | 27 | 2021-10-05 | 91 |
| 2021-01-31 | 453 | 2021-04-03 | 483 | 2021-06-04 | 245 | 2021-08-05 | 38 | 2021-10-06 | 98 |
| 2021-02-01 | 449 | 2021-04-04 | 522 | 2021-06-05 | 225 | 2021-08-06 | 43 | 2021-10-07 | 92 |
| 2021-02-02 | 462 | 2021-04-05 | 555 | 2021-06-06 | 213 | 2021-08-07 | 56 | 2021-10-08 | 93 |
| 2021-02-03 | 366 | 2021-04-06 | 580 | 2021-06-07 | 211 | 2021-08-08 | 31 | 2021-10-09 | 91 |
| 2021-02-04 | 424 | 2021-04-07 | 581 | 2021-06-08 | 220 | 2021-08-09 | 41 | 2021-10-10 | 103 |
| 2021-02-05 | 399 | 2021-04-08 | 587 | 2021-06-09 | 199 | 2021-08-10 | 34 | 2021-10-11 | 87 |
| 2021-02-06 | 399 | 2021-04-09 | 607 | 2021-06-10 | 188 | 2021-08-11 | 39 | 2021-10-12 | 86 |
| 2021-02-07 | 370 | 2021-04-10 | 630 | 2021-06-11 | 177 | 2021-08-12 | 39 | 2021-10-13 | 71 |
| 2021-02-08 | 374 | 2021-04-11 | 649 | 2021-06-12 | 164 | 2021-08-13 | 41 | 2021-10-14 | 79 |
| 2021-02-09 | 385 | 2021-04-12 | 688 | 2021-06-13 | 147 | 2021-08-14 | 42 | 2021-10-15 | 93 |
| 2021-02-10 | 367 | 2021-04-13 | 734 | 2021-06-14 | 152 | 2021-08-15 | 48 | 2021-10-16 | 80 |
| 2021-02-11 | 370 | 2021-04-14 | 745 | 2021-06-15 | 111 | 2021-08-16 | 45 | 2021-10-17 | 83 |
| 2021-02-12 | 343 | 2021-04-15 | 787 | 2021-06-16 | 104 | 2021-08-17 | 47 | 2021-10-18 | 71 |
| 2021-02-13 | 325 | 2021-04-16 | 809 | 2021-06-17 | 100 | 2021-08-18 | 48 | 2021-10-19 | 86 |
| 2021-02-14 | 296 | 2021-04-17 | 834 | 2021-06-18 | 85 | 2021-08-19 | 49 | 2021-10-20 | 85 |
| 2021-02-15 | 329 | 2021-04-18 | 874 | 2021-06-19 | 68 | 2021-08-20 | 59 | 2021-10-21 | 87 |
| 2021-02-16 | 323 | 2021-04-19 | 939 | 2021-06-20 | 63 | 2021-08-21 | 66 | 2021-10-22 | 88 |
| 2021-02-17 | 266 | 2021-04-20 | 974 | 2021-06-21 | 63 | 2021-08-22 | 74 | 2021-10-23 | 90 |
| 2021-02-18 | 324 | 2021-04-21 | 952 | 2021-06-22 | 71 | 2021-08-23 | 83 | 2021-10-24 | 84 |
| 2021-02-19 | 289 | 2021-04-22 | 961 | 2021-06-23 | 59 | 2021-08-24 | 98 | 2021-10-25 | 80 |
| 2021-02-20 | 313 | 2021-04-23 | 917 | 2021-06-24 | 57 | 2021-08-25 | 107 | 2021-10-26 | 74 |
| 2021-02-21 | 317 | 2021-04-24 | 884 | 2021-06-25 | 51 | 2021-08-26 | 118 | 2021-10-27 | 67 |
| 2021-02-22 | 296 | 2021-04-25 | 846 | 2021-06-26 | 46 | 2021-08-27 | 119 | 2021-10-28 | 62 |
| 2021-02-23 | 298 | 2021-04-26 | 877 | 2021-06-27 | 44 | 2021-08-28 | 126 | 2021-10-29 | 64 |
| 2021-02-24 | 280 | 2021-04-27 | 869 | 2021-06-28 | 49 | 2021-08-29 | 128 | 2021-10-30 | 57 |
| 2021-02-25 | 280 | 2021-04-28 | 835 | 2021-06-29 | 43 | 2021-08-30 | 129 | 2021-10-31 | 63 |
| 2021-02-26 | 273 | 2021-04-29 | 778 | 2021-06-30 | 42 | 2021-08-31 | 143 | 2021-11-01 | 69 |
| 2021-02-27 | 285 | 2021-04-30 | 735 | 2021-07-01 | 35 | 2021-09-01 | 148 | 2021-11-02 | 71 |
| 2021-02-28 | 257 | 2021-05-01 | 719 | 2021-07-02 | 36 | 2021-09-02 | 134 | 2021-11-03 | 69 |
| 2021-03-01 | 259 | 2021-05-02 | 715 | 2021-07-03 | 32 | 2021-09-03 | 131 | 2021-11-04 | 68 |
| 2021-03-02 | 261 | 2021-05-03 | 752 | 2021-07-04 | 28 | 2021-09-04 | 135 | 2021-11-05 | 61 |
| 2021-03-03 | 250 | 2021-05-04 | 736 | 2021-07-05 | 28 | 2021-09-05 | 127 | 2021-11-06 | 64 |
| 2021-11-07 | 54 | 2022-01-11 | 955 | 2022-03-17 | 117 | 2022-05-21 | 285 | 2022-07-25 | 395 |
| 2021-11-08 | 64 | 2022-01-12 | 1096 | 2022-03-18 | 102 | 2022-05-22 | 283 | 2022-07-26 | 427 |
| 2021-11-09 | 54 | 2022-01-13 | 1107 | 2022-03-19 | 111 | 2022-05-23 | 280 | 2022-07-27 | 390 |
| 2021-11-10 | 57 | 2022-01-14 | 1161 | 2022-03-20 | 104 | 2022-05-24 | 283 | 2022-07-28 | 408 |
| 2021-11-11 | 49 | 2022-01-15 | 1171 | 2022-03-21 | 116 | 2022-05-25 | 287 | 2022-07-29 | 413 |
| 2021-11-12 | 48 | 2022-01-16 | 1149 | 2022-03-22 | 130 | 2022-05-26 | 277 | 2022-07-30 | 403 |
| 2021-11-13 | 46 | 2022-01-17 | 1197 | 2022-03-23 | 143 | 2022-05-27 | 250 | 2022-07-31 | 376 |
| 2021-11-14 | 45 | 2022-01-18 | 1200 | 2022-03-24 | 152 | 2022-05-28 | 221 | 2022-08-01 | 375 |
| 2021-11-15 | 53 | 2022-01-19 | 1163 | 2022-03-25 | 155 | 2022-05-29 | 202 | 2022-08-02 | 340 |
| 2021-11-16 | 57 | 2022-01-20 | 1221 | 2022-03-26 | 155 | 2022-05-30 | 204 | 2022-08-03 | 387 |
| 2021-11-17 | 53 | 2022-01-21 | 1239 | 2022-03-27 | 145 | 2022-05-31 | 206 | 2022-08-04 | 363 |
| 2021-11-18 | 53 | 2022-01-22 | 1170 | 2022-03-28 | 163 | 2022-06-01 | 177 | 2022-08-05 | 338 |
| 2021-11-19 | 52 | 2022-01-23 | 1121 | 2022-03-29 | 173 | 2022-06-02 | 187 | 2022-08-06 | 336 |
| 2021-11-20 | 69 | 2022-01-24 | 1127 | 2022-03-30 | 177 | 2022-06-03 | 187 | 2022-08-07 | 315 |
| 2021-11-21 | 63 | 2022-01-25 | 1162 | 2022-03-31 | 184 | 2022-06-04 | 204 | 2022-08-08 | 313 |
| 2021-11-22 | 69 | 2022-01-26 | 1097 | 2022-04-01 | 172 | 2022-06-05 | 195 | 2022-08-09 | 313 |
| 2021-11-23 | 74 | 2022-01-27 | 1036 | 2022-04-02 | 202 | 2022-06-06 | 191 | 2022-08-10 | 308 |
| 2021-11-24 | 67 | 2022-01-28 | 958 | 2022-04-03 | 216 | 2022-06-07 | 174 | 2022-08-11 | 274 |
| 2021-11-25 | 59 | 2022-01-29 | 891 | 2022-04-04 | 228 | 2022-06-08 | 180 | 2022-08-12 | 288 |
| 2021-11-26 | 51 | 2022-01-30 | 820 | 2022-04-05 | 252 | 2022-06-09 | 179 | 2022-08-13 | 269 |
| 2021-11-27 | 43 | 2022-01-31 | 839 | 2022-04-06 | 252 | 2022-06-10 | 177 | 2022-08-14 | 259 |
| 2021-11-28 | 48 | 2022-02-01 | 831 | 2022-04-07 | 241 | 2022-06-11 | 170 | 2022-08-15 | 286 |
| 2021-11-29 | 59 | 2022-02-02 | 751 | 2022-04-08 | 268 | 2022-06-12 | 160 | 2022-08-16 | 300 |
| 2021-11-30 | 63 | 2022-02-03 | 706 | 2022-04-09 | 294 | 2022-06-13 | 164 | 2022-08-17 | 293 |
| 2021-12-01 | 67 | 2022-02-04 | 609 | 2022-04-10 | 270 | 2022-06-14 | 178 | 2022-08-18 | 275 |
| 2021-12-02 | 62 | 2022-02-05 | 589 | 2022-04-11 | 292 | 2022-06-15 | 169 | 2022-08-19 | 287 |
| 2021-12-03 | 69 | 2022-02-06 | 587 | 2022-04-12 | 322 | 2022-06-16 | 161 | 2022-08-20 | 301 |
| 2021-12-04 | 74 | 2022-02-07 | 567 | 2022-04-13 | 299 | 2022-06-17 | 157 | 2022-08-21 | 291 |
| 2021-12-05 | 66 | 2022-02-08 | 547 | 2022-04-14 | 314 | 2022-06-18 | 170 | 2022-08-22 | 293 |
| 2021-12-06 | 82 | 2022-02-09 | 479 | 2022-04-15 | 314 | 2022-06-19 | 150 | 2022-08-23 | 296 |
| 2021-12-07 | 91 | 2022-02-10 | 436 | 2022-04-16 | 319 | 2022-06-20 | 151 | 2022-08-24 | 306 |
| 2021-12-08 | 84 | 2022-02-11 | 388 | 2022-04-17 | 318 | 2022-06-21 | 168 | 2022-08-25 | 296 |
| 2021-12-09 | 72 | 2022-02-12 | 369 | 2022-04-18 | 320 | 2022-06-22 | 149 | 2022-08-26 | 279 |
| 2021-12-10 | 67 | 2022-02-13 | 331 | 2022-04-19 | 311 | 2022-06-23 | 159 | 2022-08-27 | 264 |
| 2021-12-11 | 76 | 2022-02-14 | 318 | 2022-04-20 | 333 | 2022-06-24 | 162 | 2022-08-28 | 261 |
| 2021-12-12 | 72 | 2022-02-15 | 322 | 2022-04-21 | 366 | 2022-06-25 | 160 | 2022-08-29 | 269 |
| 2021-12-13 | 80 | 2022-02-16 | 302 | 2022-04-22 | 368 | 2022-06-26 | 167 | 2022-08-30 | 266 |
| 2021-12-14 | 89 | 2022-02-17 | 284 | 2022-04-23 | 372 | 2022-06-27 | 189 | 2022-08-31 | 277 |
| 2021-12-15 | 61 | 2022-02-18 | 265 | 2022-04-24 | 331 | 2022-06-28 | 213 | 2022-09-01 | 267 |
| 2021-12-16 | 74 | 2022-02-19 | 237 | 2022-04-25 | 352 | 2022-06-29 | 199 | 2022-09-02 | 274 |
| 2021-12-17 | 75 | 2022-02-20 | 223 | 2022-04-26 | 342 | 2022-06-30 | 199 | 2022-09-03 | 271 |
| 2021-12-18 | 62 | 2022-02-21 | 228 | 2022-04-27 | 383 | 2022-07-01 | 171 | 2022-09-04 | 253 |
| 2021-12-19 | 62 | 2022-02-22 | 222 | 2022-04-28 | 376 | 2022-07-02 | 203 | 2022-09-05 | 277 |
| 2021-12-20 | 70 | 2022-02-23 | 210 | 2022-04-29 | 364 | 2022-07-03 | 201 | 2022-09-06 | 290 |
| 2021-12-21 | 88 | 2022-02-24 | 211 | 2022-04-30 | 370 | 2022-07-04 | 231 | 2022-09-07 | 302 |
| 2021-12-22 | 90 | 2022-02-25 | 197 | 2022-05-01 | 371 | 2022-07-05 | 247 | 2022-09-08 | 301 |
| 2021-12-23 | 103 | 2022-02-26 | 184 | 2022-05-02 | 394 | 2022-07-06 | 221 | 2022-09-09 | 306 |
| 2021-12-24 | 118 | 2022-02-27 | 161 | 2022-05-03 | 402 | 2022-07-07 | 229 | 2022-09-10 | 305 |
| 2021-12-25 | 120 | 2022-02-28 | 159 | 2022-05-04 | 373 | 2022-07-08 | 252 | 2022-09-11 | 281 |
| 2021-12-26 | 118 | 2022-03-01 | 163 | 2022-05-05 | 392 | 2022-07-09 | 252 | 2022-09-12 | 292 |
| 2021-12-27 | 131 | 2022-03-02 | 165 | 2022-05-06 | 386 | 2022-07-10 | 262 | 2022-09-13 | 315 |
| 2021-12-28 | 148 | 2022-03-03 | 167 | 2022-05-07 | 380 | 2022-07-11 | 281 | 2022-09-14 | 288 |
| 2021-12-29 | 224 | 2022-03-04 | 160 | 2022-05-08 | 373 | 2022-07-12 | 300 | 2022-09-15 | 270 |
| 2021-12-30 | 237 | 2022-03-05 | 161 | 2022-05-09 | 361 | 2022-07-13 | 309 | 2022-09-16 | 272 |
| 2021-12-31 | 327 | 2022-03-06 | 160 | 2022-05-10 | 326 | 2022-07-14 | 303 | 2022-09-17 | 282 |
| 2022-01-01 | 382 | 2022-03-07 | 164 | 2022-05-11 | 348 | 2022-07-15 | 328 | 2022-09-18 | 251 |
| 2022-01-02 | 436 | 2022-03-08 | 172 | 2022-05-12 | 347 | 2022-07-16 | 347 | 2022-09-19 | 252 |
| 2022-01-03 | 513 | 2022-03-09 | 171 | 2022-05-13 | 359 | 2022-07-17 | 367 | 2022-09-20 | 269 |
| 2022-01-04 | 591 | 2022-03-10 | 156 | 2022-05-14 | 342 | 2022-07-18 | 392 | 2022-09-21 | 258 |
| 2022-01-05 | 686 | 2022-03-11 | 145 | 2022-05-15 | 320 | 2022-07-19 | 450 | 2022-09-22 | 254 |
| 2022-01-06 | 682 | 2022-03-12 | 140 | 2022-05-16 | 319 | 2022-07-20 | 468 | 2022-09-23 | 300 |
| 2022-01-07 | 774 | 2022-03-13 | 135 | 2022-05-17 | 338 | 2022-07-21 | 464 | 2022-09-24 | 275 |
| 2022-01-08 | 766 | 2022-03-14 | 130 | 2022-05-18 | 286 | 2022-07-22 | 452 | 2022-09-25 | 251 |
| 2022-01-09 | 896 | 2022-03-15 | 108 | 2022-05-19 | 293 | 2022-07-23 | 462 | 2022-09-26 | 275 |
| 2022-01-10 | 978 | 2022-03-16 | 109 | 2022-05-20 | 293 | 2022-07-24 | 426 | 2022-09-27 | 293 |
| 2022-09-28 | 305 | 2022-10-17 | 416 | 2022-11-05 | 420 | 2022-11-24 | 322 | 2022-12-13 | 321 |
| 2022-09-29 | 301 | 2022-10-18 | 464 | 2022-11-06 | 373 | 2022-11-25 | 318 | 2022-12-14 | 277 |
| 2022-09-30 | 327 | 2022-10-19 | 434 | 2022-11-07 | 384 | 2022-11-26 | 292 | 2022-12-15 | 319 |
| 2022-10-01 | 326 | 2022-10-20 | 453 | 2022-11-08 | 385 | 2022-11-27 | 263 | 2022-12-16 | 325 |
| 2022-10-02 | 318 | 2022-10-21 | 435 | 2022-11-09 | 372 | 2022-11-28 | 267 | 2022-12-17 | 327 |
| 2022-10-03 | 315 | 2022-10-22 | 452 | 2022-11-10 | 370 | 2022-11-29 | 296 | 2022-12-18 | 313 |
| 2022-10-04 | 351 | 2022-10-23 | 422 | 2022-11-11 | 337 | 2022-11-30 | 284 | 2022-12-19 | 325 |
| 2022-10-05 | 351 | 2022-10-24 | 442 | 2022-11-12 | 325 | 2022-12-01 | 287 | 2022-12-20 | 333 |
| 2022-10-06 | 367 | 2022-10-25 | 455 | 2022-11-13 | 321 | 2022-12-02 | 279 | 2022-12-21 | 306 |
| 2022-10-07 | 393 | 2022-10-26 | 442 | 2022-11-14 | 328 | 2022-12-03 | 266 | 2022-12-22 | 300 |
| 2022-10-08 | 387 | 2022-10-27 | 436 | 2022-11-15 | 344 | 2022-12-04 | 260 | 2022-12-23 | 310 |
| 2022-10-09 | 371 | 2022-10-28 | 472 | 2022-11-16 | 312 | 2022-12-05 | 282 | 2022-12-24 | 282 |
| 2022-10-10 | 404 | 2022-10-29 | 445 | 2022-11-17 | 296 | 2022-12-06 | 288 | 2022-12-25 | 267 |
| 2022-10-11 | 418 | 2022-10-30 | 433 | 2022-11-18 | 317 | 2022-12-07 | 297 | 2022-12-26 | 267 |
| 2022-10-12 | 454 | 2022-10-31 | 445 | 2022-11-19 | 329 | 2022-12-08 | 277 | 2022-12-27 | 277 |
| 2022-10-13 | 415 | 2022-11-01 | 472 | 2022-11-20 | 307 | 2022-12-09 | 286 | 2022-12-28 | 294 |
| 2022-10-14 | 408 | 2022-11-02 | 414 | 2022-11-21 | 311 | 2022-12-10 | 308 | 2022-12-29 | 288 |
| 2022-10-15 | 446 | 2022-11-03 | 438 | 2022-11-22 | 321 | 2022-12-11 | 298 | 2022-12-30 | 297 |
| 2022-10-16 | 380 | 2022-11-04 | 407 | 2022-11-23 | 319 | 2022-12-12 | 307 | 2022-12-31 | 317 |

**Appendix B:** Data grouped into 73 groups with dates assigned numerical values

| Group | Number of Hospitalizations | Group | Number of Hospitalizations | Group | Number of Hospitalizations | Group | Number of Hospitalizations | Group | Number of Hospitalizations |
| --- | --- | --- | --- | --- | --- | --- | --- | --- | --- |
| 1 | 5664 | 16 | 2457 | 31 | 648 | 46 | 1912 | 61 | 2759 |
| 2 | 6520 | 17 | 1296 | 32 | 527 | 47 | 2866 | 62 | 2878 |
| 3 | 5278 | 18 | 546 | 33 | 610 | 48 | 3390 | 63 | 2711 |
| 4 | 4081 | 19 | 329 | 34 | 633 | 49 | 3767 | 64 | 2971 |
| 5 | 3232 | 20 | 309 | 35 | 766 | 50 | 3542 | 65 | 3811 |
| 6 | 2858 | 21 | 324 | 36 | 906 | 51 | 2947 | 66 | 4303 |
| 7 | 2493 | 22 | 341 | 37 | 3675 | 52 | 2115 | 67 | 4464 |
| 8 | 2723 | 23 | 424 | 38 | 9586 | 53 | 1768 | 68 | 3900 |
| 9 | 3712 | 24 | 899 | 39 | 11749 | 54 | 1596 | 69 | 3190 |
| 10 | 5481 | 25 | 1343 | 40 | 8538 | 55 | 1933 | 70 | 2969 |
| 11 | 8033 | 26 | 1302 | 41 | 4611 | 56 | 2656 | 71 | 2841 |
| 12 | 8654 | 27 | 1057 | 42 | 2504 | 57 | 4156 | 72 | 3153 |
| 13 | 6630 | 28 | 884 | 43 | 1677 | 58 | 3914 | 73 | 2899 |
| 14 | 5235 | 29 | 866 | 44 | 1430 | 59 | 3117 |  |  |
| 15 | 3979 | 30 | 812 | 45 | 1285 | 60 | 2881 |  |  |

**Appendix C:** Number of hospitalizations in increasing order and with term numbers

| Term | Number of Hospitalizations | Term | Number of Hospitalizations | Term | Number of Hospitalizations | Term | Number of Hospitalizations | Term | Number of Hospitalizations |
| --- | --- | --- | --- | --- | --- | --- | --- | --- | --- |
| 1 | 309 | 16 | 906 | 31 | 2504 | 46 | 3153 | 61 | 4464 |
| 2 | 324 | 17 | 1057 | 32 | 2656 | 47 | 3190 | 62 | 4611 |
| 3 | 329 | 18 | 1285 | 33 | 2711 | 48 | 3232 | 63 | 5235 |
| 4 | 341 | 19 | 1296 | 34 | 2723 | 49 | 3390 | 64 | 5278 |
| 5 | 424 | 20 | 1302 | 35 | 2759 | 50 | 3542 | 65 | 5481 |
| 6 | 527 | 21 | 1343 | 36 | 2841 | 51 | 3675 | 66 | 5664 |
| 7 | 546 | 22 | 1430 | 37 | 2858 | 52 | 3712 | 67 | 6520 |
| 8 | 610 | 23 | 1596 | 38 | 2866 | 53 | 3767 | 68 | 6630 |
| 9 | 633 | 24 | 1677 | 39 | 2878 | 54 | 3811 | 69 | 8033 |
| 10 | 648 | 25 | 1768 | 40 | 2881 | 55 | 3900 | 70 | 8538 |
| 11 | 766 | 26 | 1912 | 41 | 2899 | 56 | 3914 | 71 | 8654 |
| 12 | 812 | 27 | 1933 | 42 | 2947 | 57 | 3979 | 72 | 9586 |
| 13 | 866 | 28 | 2115 | 43 | 2969 | 58 | 4081 | 73 | 11749 |
| 14 | 884 | 29 | 2457 | 44 | 2971 | 59 | 4156 |  |  |
| 15 | 899 | 30 | 2493 | 45 | 3117 | 60 | 4303 |  |  |

**Appendix D:** Data grouped into 73 groups with dates assigned numerical values and outliers replaced by the mean of the two closest number of hospitalizations

| Group | Number of Hospitalizations | Group | Number of Hospitalizations | Group | Number of Hospitalizations | Group | Number of Hospitalizations | Group | Number of Hospitalizations |
| --- | --- | --- | --- | --- | --- | --- | --- | --- | --- |
| 1 | 5664 | 16 | 2457 | 31 | 648 | 46 | 1912 | 61 | 2759 |
| 2 | 6520 | 17 | 1296 | 32 | 527 | 47 | 2866 | 62 | 2878 |
| 3 | 5278 | 18 | 546 | 33 | 610 | 48 | 3390 | 63 | 2711 |
| 4 | 4081 | 19 | 329 | 34 | 633 | 49 | 3767 | 64 | 2971 |
| 5 | 3232 | 20 | 309 | 35 | 766 | 50 | 3542 | 65 | 3811 |
| 6 | 2858 | 21 | 324 | 36 | 906 | 51 | 2947 | 66 | 4303 |
| 7 | 2493 | 22 | 341 | 37 | 3675 | 52 | 2115 | 67 | 4464 |
| 8 | 2723 | 23 | 424 | 38 | 4143 | 53 | 1768 | 68 | 3900 |
| 9 | 3712 | 24 | 899 | 39 | 4143 | 54 | 1596 | 69 | 3190 |
| 10 | 5481 | 25 | 1343 | 40 | 4143 | 55 | 1933 | 70 | 2969 |
| 11 | 6055.5 | 26 | 1302 | 41 | 4611 | 56 | 2656 | 71 | 2841 |
| 12 | 6055.5 | 27 | 1057 | 42 | 2504 | 57 | 4156 | 72 | 3153 |
| 13 | 6630 | 28 | 884 | 43 | 1677 | 58 | 3914 | 73 | 2899 |
| 14 | 5235 | 29 | 866 | 44 | 1430 | 59 | 3117 |  |  |
| 15 | 3979 | 30 | 812 | 45 | 1285 | 60 | 2881 |  |  |

**Appendix E:** Link to the Code Randomizing Fifteen Numbers from 1 to 73 https://docs.google.com/document/d/1iTKYf4wEY5faM6ikTFJ7JEy7ghZVIxSkIOOK15JgwPo/edit?usp=sharing

**Appendix F:** Data Grouped into 73 Groups with Dates Assigned Numerical Values, with Outliers Replaced, Only Including the Train Split

| Group | Number of Hospitalizations | Group | Number of Hospitalizations | Group | Number of Hospitalizations | Group | Number of Hospitalizations |
| --- | --- | --- | --- | --- | --- | --- | --- |
| 1 | 5664 | 19 | 329 | 38 | 4143 | 57 | 4156 |
| 2 | 6520 | 20 | 309 | 39 | 4143 | 58 | 3914 |
| 3 | 5278 | 21 | 324 | 40 | 4143 | 59 | 3117 |
| 4 | 4081 | 22 | 341 | 41 | 4611 | 60 | 2881 |
| 6 | 2858 | 23 | 424 | 42 | 2504 | 61 | 2759 |
| 7 | 2493 | 24 | 899 | 44 | 1430 | 62 | 2878 |
| 9 | 3712 | 25 | 1343 | 45 | 1285 | 63 | 2711 |
| 10 | 5481 | 27 | 1057 | 47 | 2866 | 65 | 3811 |
| 11 | 6055.5 | 28 | 884 | 48 | 3390 | 66 | 4303 |
| 12 | 6055.5 | 29 | 866 | 49 | 3767 | 67 | 4464 |
| 13 | 6630 | 31 | 648 | 50 | 3542 | 68 | 3900 |
| 14 | 5235 | 32 | 527 | 52 | 2115 | 70 | 2969 |
| 15 | 3979 | 33 | 610 | 53 | 1768 | 73 | 2899 |
| 16 | 2457 | 34 | 633 | 54 | 1596 |  |  |
| 18 | 546 | 36 | 906 | 55 | 1933 |  |  |

## Footnotes

1 Meisa Salaita, “10 Ways We’Re Using Data to Fight Disease,” HowStuffWorks, August 20, 2020, https://science.howstuffworks.com/life/genetic/10-ways-were-using-data-fight-disease.htm.

2 “COVID-19 Cases in Hospital and ICU, by Ontario Health (OH) Region - Ontario Data Catalogue,” n.d., https://data.ontario.ca/dataset/covid-19-cases-in-hospital-and-icu-by-ontario-health-region.

3 Mark LeBoeuf, “Time Series Outlier Detection,” The Code Forest, July 29, 2017, https://thecodeforest.github.io/post/time_series_outlier_detection.html.

4 Michael Galarnyk, “Understanding Train Test Split,” Built In, July 28, 2022, https://builtin.com/data-science/train-test-split.

5 Conor Mack, “Machine Learning Fundamentals (I): Cost Functions and Gradient Descent,” Medium, April 4, 2021, https://towardsdatascience.com/machine-learning-fundamentals-via-linear-regression-41a5d11f5220.

6 Jason Brownlee, “Train-Test Split for Evaluating Machine Learning Algorithms,” Machine Learning Mastery, August 26, 2020, https://machinelearningmastery.com/train-test-split-for-evaluating-machine-learning-algorithms/.

7 Wallstreetmojo Team, “Residual Sum of Squares,” WallStreetMojo, June 18, 2022, https://www.wallstreetmojo.com/residual-sum-of-squares/.

